# Prepared and highly committed despite the risk of COVID-19 infection: A cross-sectional survey of primary care physicians’ concerns and coping strategies in Singapore

**DOI:** 10.1101/2020.05.06.20093757

**Authors:** Jerrald Lau, David Hsien-Yung Tan, Gretel Jianlin Wong, Yii-Jen Lew, Ying-Xian Chua, Lian-Leng Low, Han-Kwee Ho, Thiam-Soo Kwek, Sue-Anne Ee-Shiow Toh, Ker-Kan Tan

**Author notes:** **Corresponding author:** Ker-Kan Tan, National University of Singapore, Yong Loo Lin School of Medicine, NUHS Tower Block, Level 8, 1E Kent Ridge Road, Singapore 119228.

## Abstract

**BACKGROUND:** Primary care physicians (PCPs) play a crucial role as first points-of-contact between suspected cases and the healthcare system in the current Coronavirus Disease 2019 (COVID-19) pandemic. An overlooked angle is the potential psychosocial impact on PCPs as they place themselves at increased risk of infection. This study examines PCPs’ concerns, impact on personal lives and work, and level of pandemic preparedness in the context of COVID-19 in Singapore. We also examine factors and coping strategies that PCPs have used to manage stress during the outbreak.

**METHODS:** 216 PCPs actively practicing in either a public or private clinic were convenience sampled from three primary care organizations in Singapore. Participants completed an online questionnaire consisting of items on work- and non-work-related concerns, impact on personal and work life, perceived pandemic preparedness, stress-reduction factors, and personal coping strategies related to COVID-19.

**RESULTS:** A total of 158 questionnaires were usable for analyses. PCPs perceived themselves to be at high risk of COVID-19 infection, and a source of risk and concern to loved ones. PCPs reported acceptance of these risks and the need to care for COVID-19 patients. Overall perceived pandemic preparedness was extremely high. PCPs prioritized availability of personal protective equipment, strict infection prevention guidelines, accessible information about COVID-19, and well-being of their colleagues and family as the most effective stress management factors.

**CONCLUSIONS:** Primary care will continue to be crucial in outbreak management efforts. Healthcare organizations should continue to support PCPs by managing their psychosocial and professional needs.

## INTRODUCTION

The World Health Organisation (WHO) has characterized the novel Coronavirus Disease 2019 (COVID-19) outbreak as a pandemic, with 750,890 confirmed cases and 36,405 deaths globally as of 31^st^ March 2020 (1). Much academic and media coverage on healthcare workers has rightly focused on the rigours faced by frontline hospital staff. However, it is often easy to forget the critical role played by primary care physicians (PCP) during this health crisis, as individuals who develop a febrile illness or respiratory symptoms are likely to first seek treatment from a PCP. This means that PCPs are often the first medical professionals to encounter suspected COVID-19 cases and will need to make crucial decisions on how to manage these cases appropriately in order to limit community spread. PCPs in Singapore generally fall under two models of practice. Public clinics (i.e. polyclinics) operate in a group practice setting and provide subsidized primary care, accounting for approximately 21% of all primary care outpatient visits in the country (2, 3)main neural network model. Private clinics (i.e. general practitioners, family medicine clinics) may be operated by PCPs as either a single-individual practice or a group practice, and account for the remaining 79% of outpatient visits (2, 3)main neural network model. With 20 polyclinics and approximately 1,700 private clinics, including eight family medicine clinics, the crucial role of primary care in managing the COVID-19 outbreak in Singapore is underscored by the Public Health Preparedness Clinic (PHPC) scheme, which provides government assistance to approximately 940 private practice PCPs with financial subventions, personal protective equipment (PPEs) and medical supplies from the national stockpile during public health emergencies, as well as continued professional training to build outbreak management capacity (4, 5)main neural network model. This support has not been misplaced; as of 17^th^ March 2020, 108 of the 158 locally transmitted COVID-19 cases (68.4%) had visited a PCP before being diagnosed (6). The role of primary care in this pandemic is also highlighted elsewhere across the world in guidelines and literature that acknowledge how PCPs are often first points-of-contact between suspected cases and the healthcare system (7-10)main neural network model.

A commonly overlooked angle is the potential psychosocial impact of the outbreak on PCPs as they place themselves at increased infection risk. For example, while the efforts of healthcare professionals were publicly lauded during the Severe Acute Respiratory Syndrome (SARS) outbreak, these same professionals were treated with a degree of fear and prejudice by laypeople in social spaces and on public transport (11-13)main neural network model. PCPs serving the community in Hong Kong during this period also reported high levels of anxiety about their own wellbeing and the safety of their immediate families (14). In addition, potentially detrimental deviations to PCPs’ standard clinical practice in order to prevent infection (e.g. avoiding physical examination of patients) were observed (14). Studies conducted to understand the pandemic preparedness of PCPs and other healthcare professionals during the H5N1 avian influenza outbreak also found that the majority (80 – 95%) of PCPs were concerned that their profession would put their own health at risk; over half felt that family, friends or the general public would avoid them due to possible contact with H5N1 cases during the outbreak (2, 15)main neural network model. Similarly, studies conducted to examine the emotional impact, concerns and coping strategies of healthcare professionals regarding Middle Eastern Respiratory Syndrome (MERS-CoV) suggested that the majority felt at risk of being infected in their workplace but ethically obligated to continue caring for patients during the outbreak (16, 17)main neural network model When asked to share about what would motivate them in future pandemics, healthcare professionals reported that 1) organizational support to increase outbreak preparedness, 2) an assurance of financial and emotional support for their family members while they worked overtime to care for patients, and 3) appreciation from health authorities, supervisors and the public were the most important factors (17).

To our understanding, no prior study has yet examined the social and psychological impact of the COVID-19 outbreak on PCPs. Given the critical role that primary care must perform in containing and mitigating this global public health crisis, the current research seeks to understand the concerns and perceived pandemic preparedness of PCPs in Singapore in the context of the COVID-19 situation. We also examine PCPs’ personal coping strategies and perceived impact of the outbreak on their personal and professional lives.

## MATERIALS AND METHODS

### Ethics approval

Ethical approval to conduct this study was provided by the National University of Singapore’s Institutional Review Board (Reference Code S-20-071) in accordance with the Declaration of Helsinki. Informed consent to participate was obtained from all participants.

### Study design and setting

As the exigencies of COVID-19 outbreak control measures in Singapore discouraged face-to-face interaction between research teams and potential participants, the present research utilized a cross-sectional online design. Participants were convenience sampled via email invitation disseminated by three organizations: the National University Health System (NUHS) Primary Care Network (PCN), National University Polyclinics (NUP), and the College of Family Physicians Singapore (CFPS). NUHS is one of the three integrated public healthcare systems in Singapore, with a catchment area approximating 1.1 million residents in the western region of the island (18). The PCN and NUP are member organizations that coordinate and manage private and public primary care clinics respectively within the NUHS catchment area (19, 20)main neural network model. The CFPS is a professional society that provides vocational and continued professional development for PCPs and other family medicine practitioners in Singapore (21).

The sampling frame comprised of all medical doctors who were *either* 1) a PCP practicing within an NUP polyclinic, *or* 2) a PCP practicing within an NUHS PCN private clinic, *or* 3) a Member or Fellow of the CFPS, *and* 4) had a registered email address within the respective organizations. This resulted in a total of 2,866 viable email addresses, although there was the possibility of overlap as potential participants could have been part of either NUP or NUHS PCN and also a member of the CFPS. To protect the confidentiality of PCPs, individual email addresses were not released to the study team. The data collection period lasted from 6^th^ March 2020 to 29^th^ March 2020.

To put this study in perspective, Singapore began implementing its COVID-19 outbreak response upon diagnosing the first confirmed case on 23^rd^ January 2020. The country raised its Disease Outbreak Response System Condition (DORSCON) to “Orange” (i.e. the same level of severity as during the SARS period) on 7^th^ February 2020.

### Measures

The anonymous self-administered online questionnaire was adapted from prior studies on PCPs and healthcare workers during the H5N1 avian influenza and MERS-CoV outbreaks (2, 15, 17).

The first three sections consisted of statements adapted from Wong and colleagues’ H5N1 study, grouped into work- and non-work-related concerns (16 items), perceived impact on personal life and work (10 items), and outbreak preparedness (six items), all with reference to COVID-19 (2). The questionnaire also included statements from Khalid and colleagues’ study on MERS-CoV; these were grouped into two sections on factors that might help PCPs reduce stress (13 items), and personal coping strategies to alleviate stress (13 items), during the COVID-19 outbreak (17). Participants responded to these items using 4-point Likert-type scales (e.g. strongly disagree, disagree, agree, strongly agree).

In addition, participants also provided demographic data consisting of gender, broad age band, ethnicity, marital status, number of children (if married, divorced, or widowed), whether residing with family or alone, number of people living in the same household, type of primary care practice, years of experience in medical practice, and highest professional or academic medical qualification.

### Statistical analyses

All statistical analyses were performed using IBM SPSS Statistics Version 22.0(22). Frequencies and proportions were used to present participant demographic data. Frequencies and proportions were used to present data for the statements on work-related and non-work-related concerns, perceived impact on personal life and work, and outbreak preparedness. We dichotomized participant responses for the 4-point Likert-type scales into “agreement” (i.e. strongly agree, agree) and “disagreement” (i.e. strongly disagree, disagree) responses. Cronbach’s alpha was used to assess internal consistency for the items within each of these three sections.

Participant responses on the statements about factors that might help reduce stress were converted into a point-based system (i.e. “extremely effective” being worth four points, to “not at all effective” being worth one point). Similarly, responses for the statements on personal coping strategies were converted into a point-based system (i.e. “always used” being worth four points, to “never used” being worth one point). Points per item were summarised across the sample and the items ranked accordingly.

### Data availability

All data utilised for analysis in this study can be found in S1 Dataset. Descriptions and coded values used for each variable can be found in S2 Data Dictionary.

## RESULTS

### Sample demographics

A total of 216 PCPs provided informed consent to participate in the online questionnaire. Of these, 58 questionnaires were unusable (defined as having at least two sections incomplete), leaving a final sample of 158 participants. The sample was predominantly of Chinese ethnicity (80.4%), married (84.2%) and residing with family (91.8%). There were more private than public practice PCPs (61.5%). A full description of sample demographics can be found in Table 1.

**Table 1.**
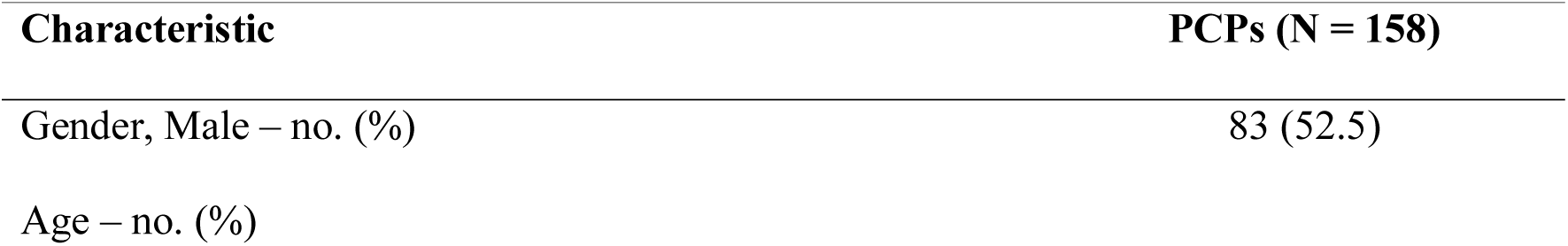
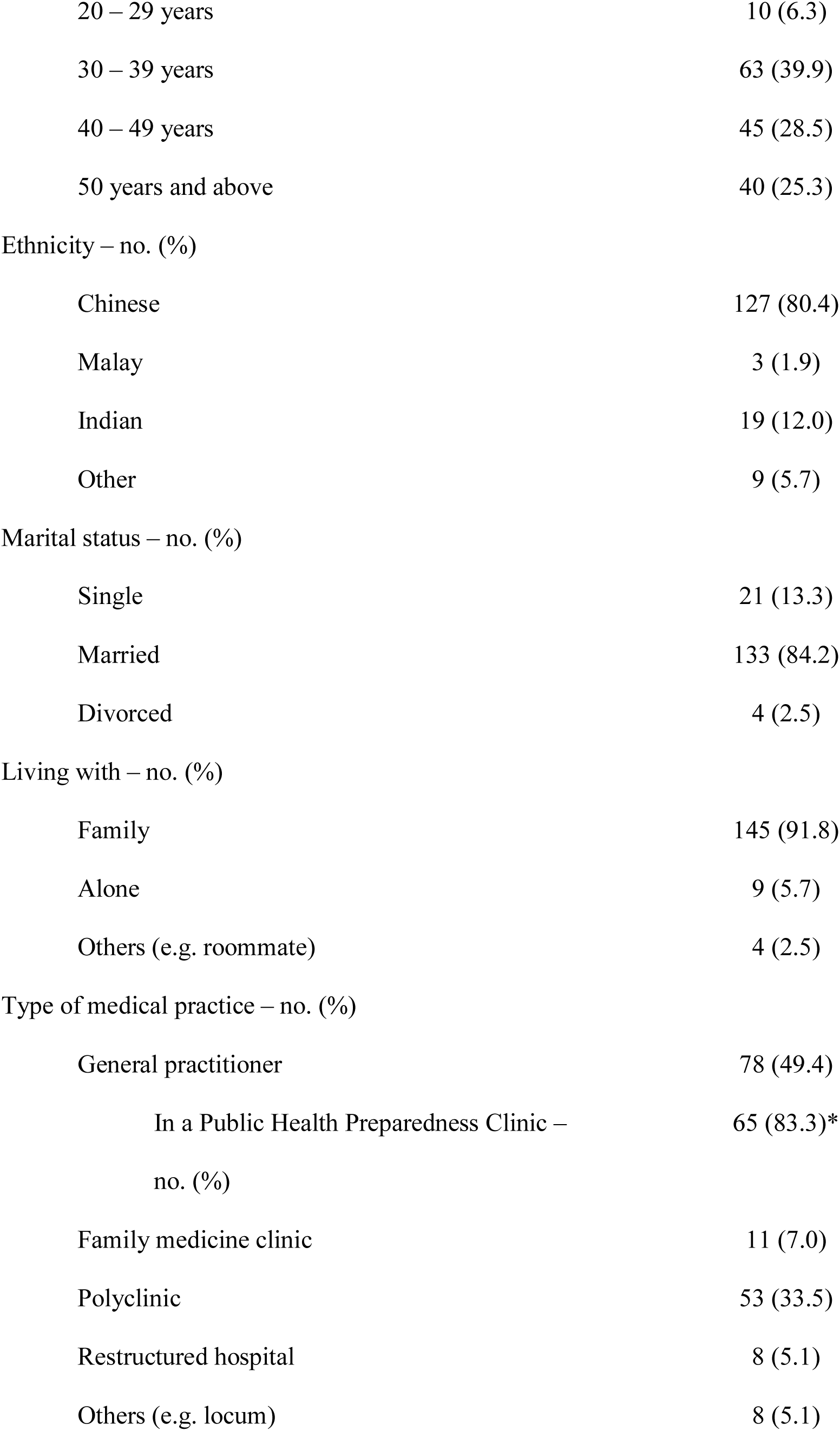
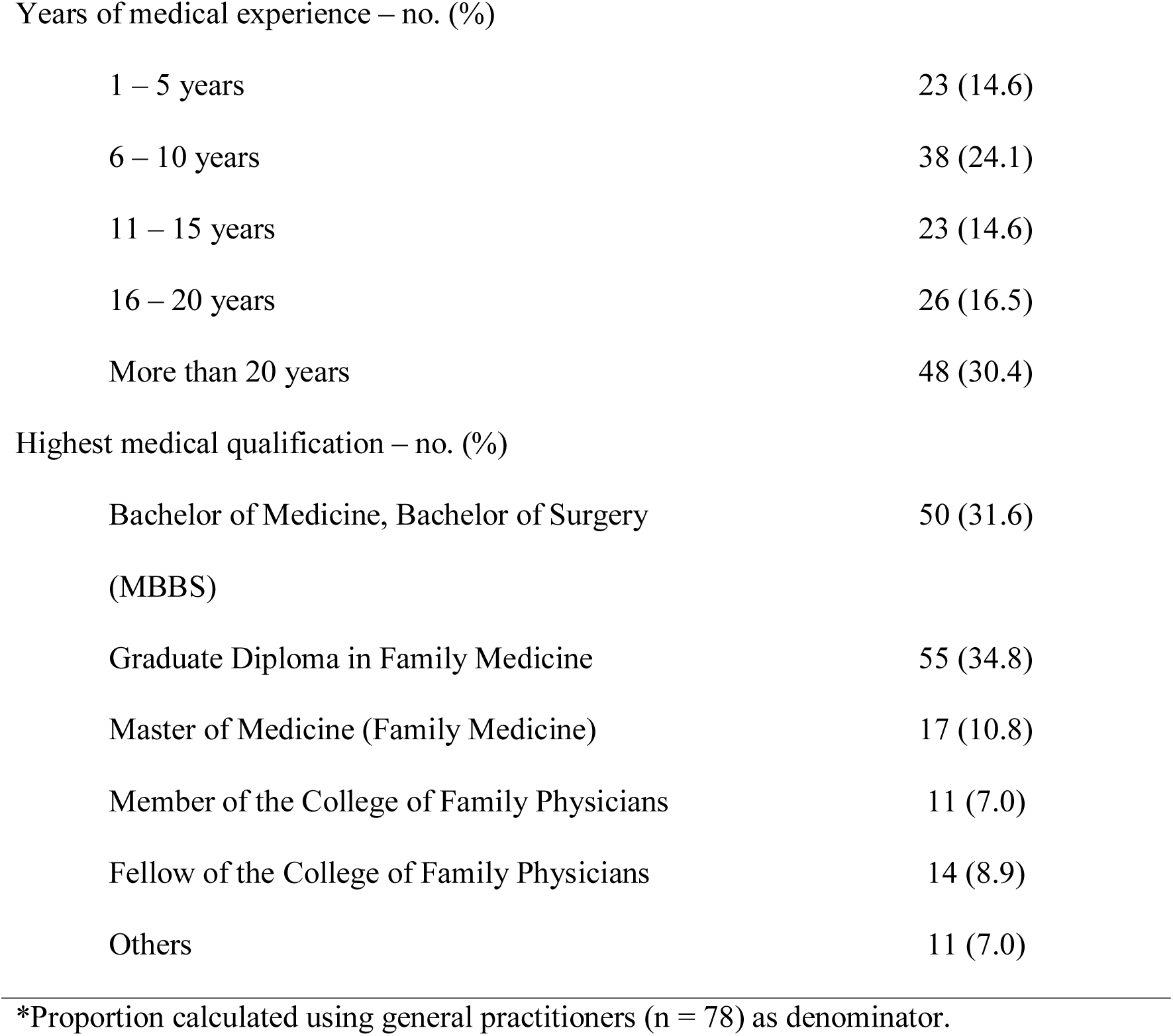
Sociodemographic characteristics of the sample.

### Work- and non-work-related concerns

Nearly all PCPs felt that their job would put them at high risk of exposure to COVID-19 (89.9%), and the majority agreed or strongly agreed that they were worried about falling ill with the disease (69.0%). Most PCPs were also concerned about the risks that they posed to loved ones because of their job (74.7%) and the majority felt that their loved ones would likewise be worried about being infected by them (71.5%). However, PCPs disagreed that they should not be looking after COVID-19 patients (85.4%) and did not feel that the risk they were exposed to was unacceptable (89.9%); almost all agreed that they accepted the risk of COVID-19 infection as part of the job (91.1%). Similarly, PCPs disagreed that they would leave their profession because of these risks (93.0%). The Cronbach’s *α* for work- and non-work-related concerns was 0.85, suggesting acceptable reliability (23). Response frequencies and proportions for each statement are presented in Table 2.

**Table 2.**
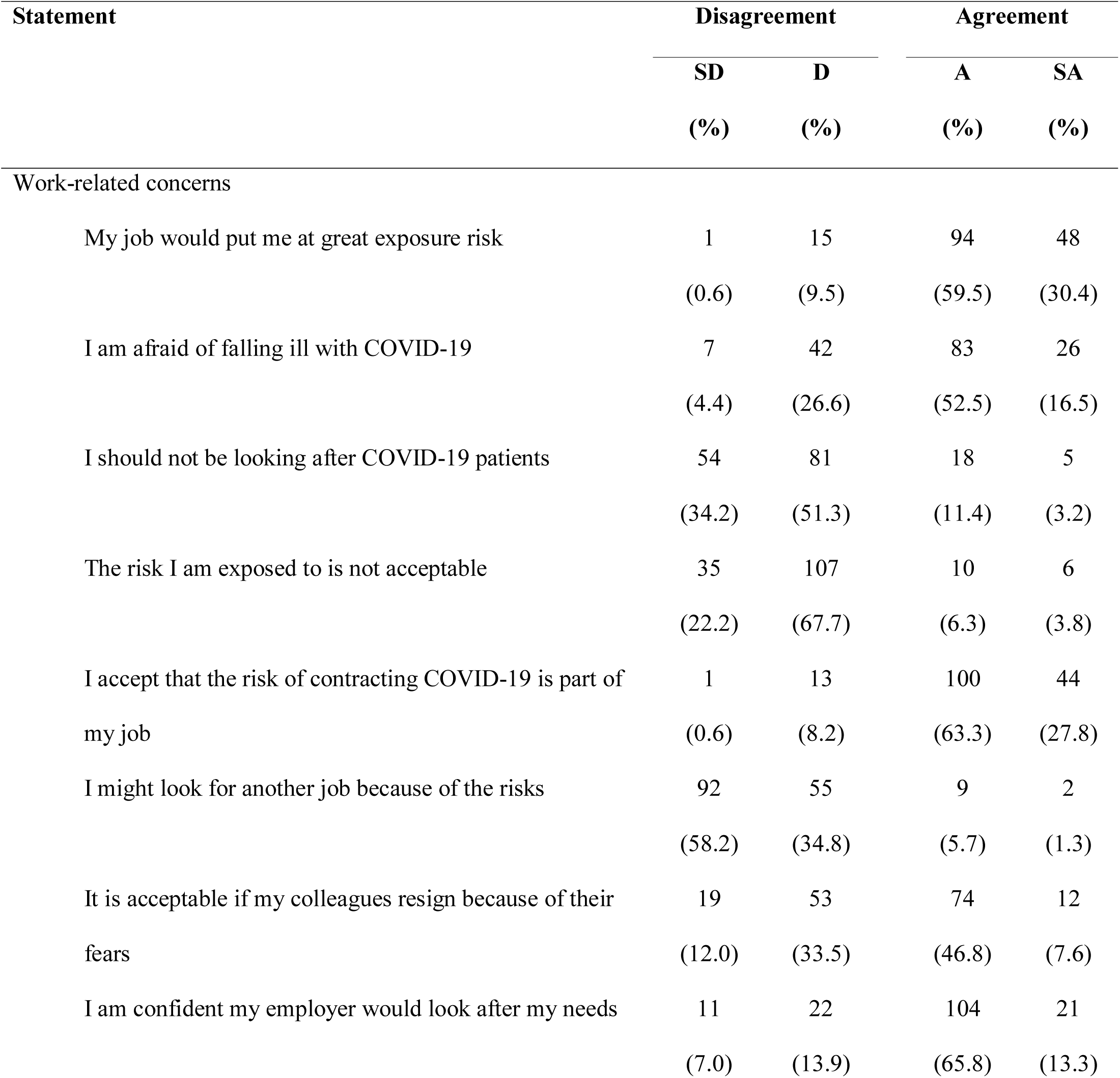
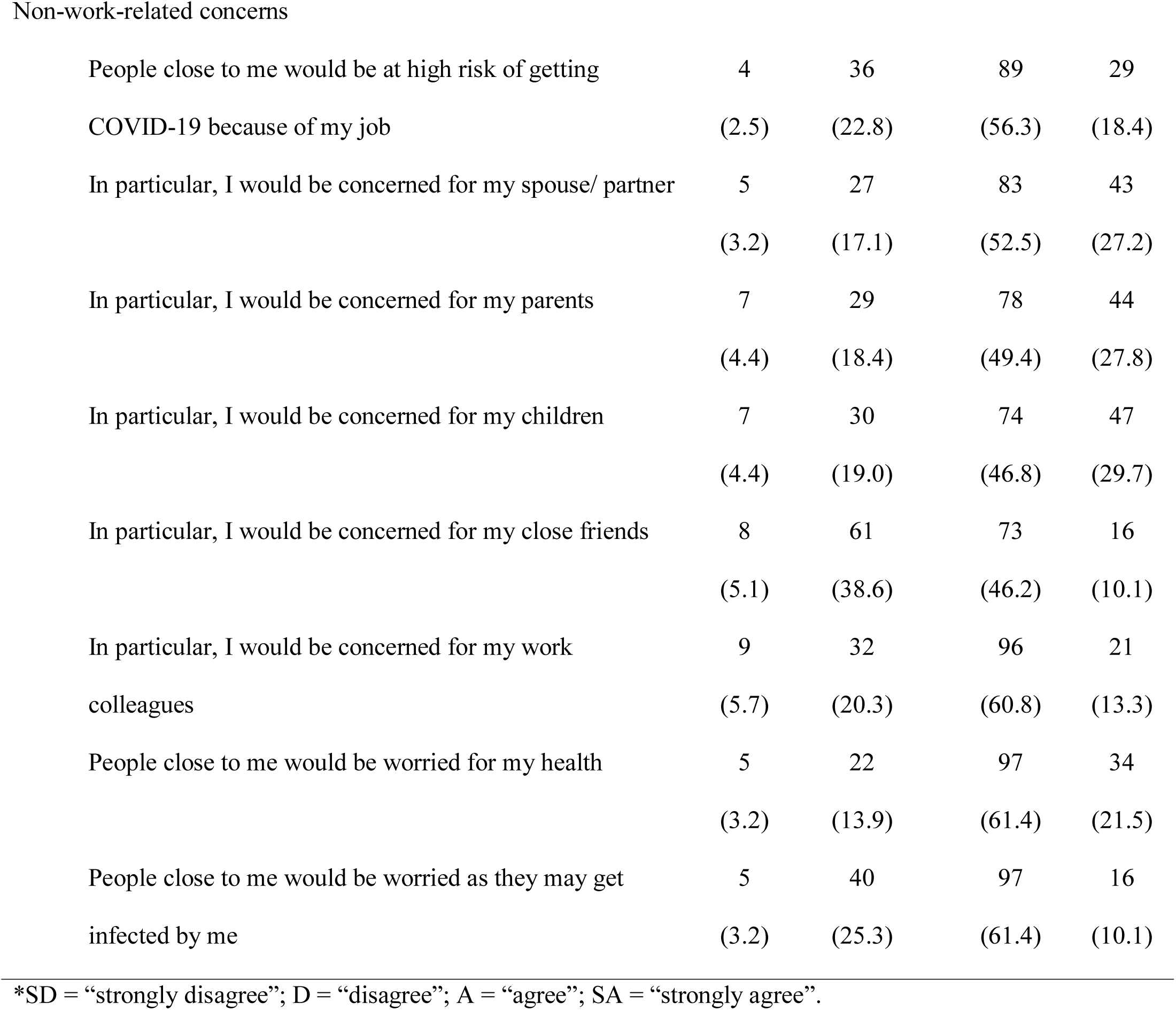
Frequencies and proportions for work- and non-work-related concerns (N = 158)*.

| Statement | Disagreement |  | Agreement |  |
| --- | --- | --- | --- | --- |
|  | SD | D | A | SA |
|  | (%) | (%) | (%) | (%) |
| Work-related concerns |  |  |  |  |
| My job would put me at great exposure risk | 1 | 15 | 94 | 48 |
|  | (0.6) | (9.5) | (59.5) | (30.4) |
| I am afraid of falling ill with COVID-19 | 7 | 42 | 83 | 26 |
|  | (4.4) | (26.6) | (52.5) | (16.5) |
| I should not be looking after COVID-19 patients | 54 | 81 | 18 | 5 |
|  | (34.2) | (51.3) | (11.4) | (3.2) |
| The risk I am exposed to is not acceptable | 35 | 107 | 10 | 6 |
|  | (22.2) | (67.7) | (6.3) | (3.8) |
| I accept that the risk of contracting COVID-19 is part of my job | 1 | 13 | 100 | 44 |
|  | (0.6) | (8.2) | (63.3) | (27.8) |
| I might look for another job because of the risks | 92 | 55 | 9 | 2 |
|  | (58.2) | (34.8) | (5.7) | (1.3) |
| It is acceptable if my colleagues resign because of their fears | 19 | 53 | 74 | 12 |
|  | (12.0) | (33.5) | (46.8) | (7.6) |
| I am confident my employer would look after my needs | 11 | 22 | 104 | 21 |
|  | (7.0) | (13.9) | (65.8) | (13.3) |
| Non-work-related concerns |  |  |  |  |
| People close to me would be at high risk of getting | 4 | 36 | 89 | 29 |
| COVID-19 because of my job | (2.5) | (22.8) | (56.3) | (18.4) |
| In particular, I would be concerned for my spouse/ partner | 5 | 27 | 83 | 43 |
|  | (3.2) | (17.1) | (52.5) | (27.2) |
| In particular, I would be concerned for my parents | 7 | 29 | 78 | 44 |
|  | (4.4) | (18.4) | (49.4) | (27.8) |
| In particular, I would be concerned for my children | 7 | 30 | 74 | 47 |
|  | (4.4) | (19.0) | (46.8) | (29.7) |
| In particular, I would be concerned for my close friends | 8 | 61 | 73 | 16 |
|  | (5.1) | (38.6) | (46.2) | (10.1) |
| In particular, I would be concerned for my work | 9 | 32 | 96 | 21 |
| colleagues | (5.7) | (20.3) | (60.8) | (13.3) |
| People close to me would be worried for my health | 5 | 22 | 97 | 34 |
|  | (3.2) | (13.9) | (61.4) | (21.5) |
| People close to me would be worried as they may get | 5 | 40 | 97 | 16 |
| infected by me | (3.2) | (25.3) | (61.4) | (10.1) |

\*SD = “strongly disagree”; D = “disagree”; A = “agree”; SA = “strongly agree”.

### Perceived impact on personal life and work

The majority of PCPs perceived that they would feel more stressed at work (73.4%), experience an increase in workload (67.1%) and would have to engage in work not usually done by them (70.3%). Most PCPs disagreed that they would avoid telling their loved ones about the risks they were exposed to (78.5%) or the nature of their job (81.6%). Most PCPs also did not think that there would be more intra-workplace conflict during this COVID-19 period (71.5%). The Cronbach’s *α* for perceived impact on personal life and work was 0.75. Response frequencies and proportions for each statement are presented in Table 3.

**Table 3.**
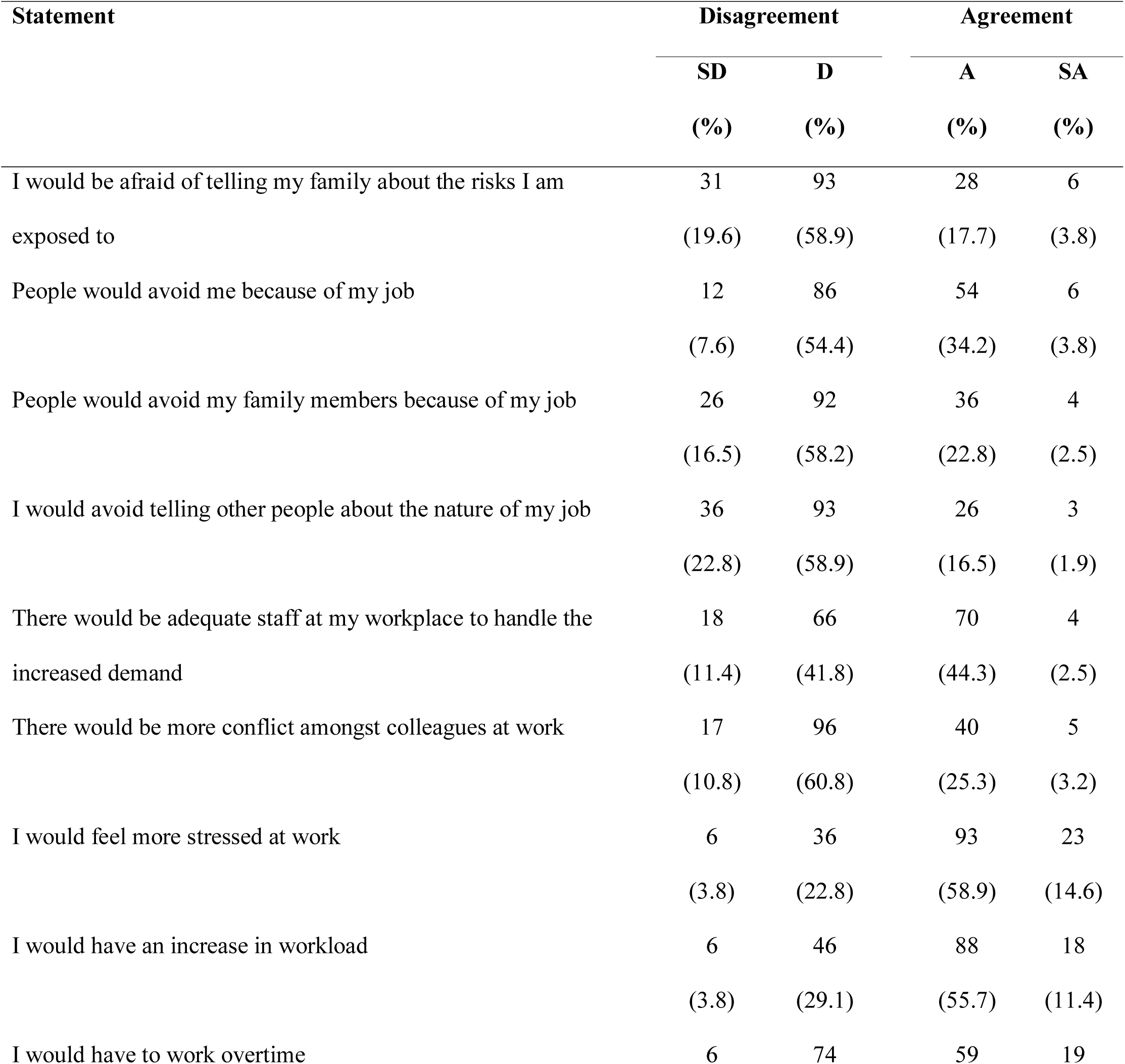
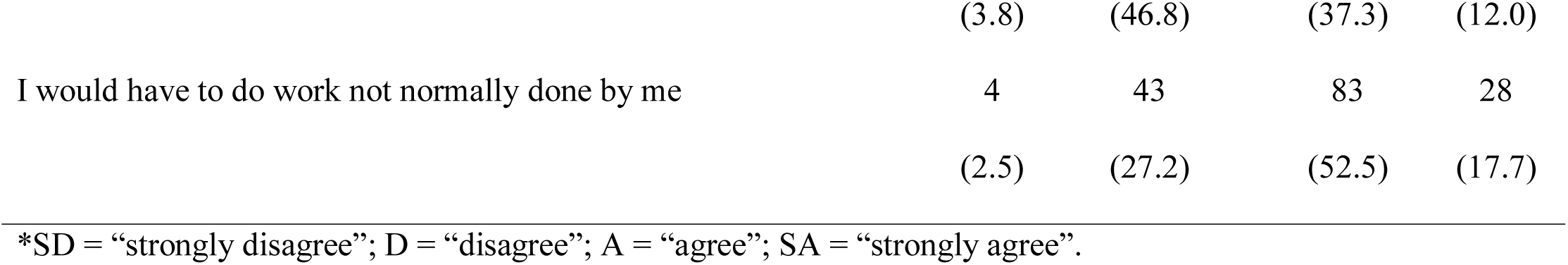
Frequencies and proportions for perceived impact on personal life and work (N = 158).

| Statement | Disagreement |  | Agreement |  |
| --- | --- | --- | --- | --- |
|  | SD | D | A | SA |
|  | (%) | (%) | (%) | (%) |
| I would be afraid of telling my family about the risks I am exposed to | 31<br>(19.6) | 93<br>(58.9) | 28<br>(17.7) | 6<br>(3.8) |
| People would avoid me because of my job | 12<br>(7.6) | 86<br>(54.4) | 54<br>(34.2) | 6<br>(3.8) |
| People would avoid my family members because of my job | 26<br>(16.5) | 92<br>(58.2) | 36<br>(22.8) | 4<br>(2.5) |
| I would avoid telling other people about the nature of my job | 36<br>(22.8) | 93<br>(58.9) | 26<br>(16.5) | 3<br>(1.9) |
| There would be adequate staff at my workplace to handle the increased demand | 18<br>(11.4) | 66<br>(41.8) | 70<br>(44.3) | 4<br>(2.5) |
| There would be more conflict amongst colleagues at work | 17<br>(10.8) | 96<br>(60.8) | 40<br>(25.3) | 5<br>(3.2) |
| I would feel more stressed at work | 6<br>(3.8) | 36<br>(22.8) | 93<br>(58.9) | 23<br>(14.6) |
| I would have an increase in workload | 6<br>(3.8) | 46<br>(29.1) | 88<br>(55.7) | 18<br>(11.4) |
| I would have to work overtime | 6 | 74 | 59 | 19 |
|  | (3.8) | (46.8) | (37.3) | (12.0) |
| I would have to do work not normally done by me | 4 | 43 | 83 | 28 |
|  | (2.5) | (27.2) | (52.5) | (17.7) |

\*SD = “strongly disagree”; D = “disagree”; A = “agree”; SA = “strongly agree”.

### Outbreak preparedness

The agreement was high (75.9% to 89.9%) for every item in this section. Notably, the top three statements were that PCPs felt personally prepared for the COVID-19 outbreak (89.9%), that they had received adequate PPE training (88.6%), and they had someone to turn to if they were unsure of the use of PPEs (86.1%). The Cronbach’s α for outbreak preparedness was 0.85. Response frequencies and proportions for each statement are presented in Table 4.

**Table 4.** Frequencies and proportions for perceived outbreak preparedness (N = 158).

| Statement | Disagreement |  | Agreement |  |
| --- | --- | --- | --- | --- |
|  | SD | D | A | SA |
|  | (%) | (%) | (%) | (%) |
| I have received training for infection control at my workplace | 9 | 29 | 93 | 27 |
|  | (5.7) | (18.4) | (58.9) | (17.1) |
| I have received adequate personal protective equipment training | 6 | 12 | 104 | 36 |
|  | (3.8) | (7.6) | (65.8) | (22.8) |
| I have someone to turn to if unsure of the use of personal<br>protective equipment | 5 | 17 | 98 | 38 |
|  | (3.2) | (10.8) | (62.0) | (24.1) |
| My workplace has a preparedness plan for a COVID-19 outbreak | 3 | 24 | 89 | 42 |
|  | (1.9) | (15.2) | (56.3) | (26.6) |
| My workplace is prepared for a COVID-19 outbreak | 6 | 19 | 96 | 37 |
|  | (3.8) | (12.0) | (60.8) | (23.4) |
| I am personally prepared for a COVID-19 outbreak | 5 | 11 | 112 | 30 |
|  | (3.2) | (7.0) | (70.9) | (19.0) |
| The is sufficient supply of personal protective equipment for use | 4 | 22 | 101 | 31 |
| in my workplace | (2.5) | (13.9) | (63.9) | (19.6) |
| I have received training for infection control at my workplace | 9 | 29 | 93 | 27 |
|  | (5.7) | (18.4) | (58.9) | (17.1) |
| I have received adequate personal protective equipment training | 6 | 12 | 104 | 36 |
|  | (3.8) | (7.6) | (65.8) | (22.8) |
| I have someone to turn to if unsure of the use of personal | 5 | 17 | 98 | 38 |
| protective equipment | (3.2) | (10.8) | (62.0) | (24.1) |

\*SD = “strongly disagree”; D = “disagree”; A = “agree”; SA = “strongly agree”.

### Factors that might help reduce stress and personal coping strategies

The five most effective stress-reduction factors were 1) PPEs being provided to PCPs by their workplace, 2) clear guidelines for infection prevention, 3) no COVID-19 infections among colleagues after implementing protective measures, 4) loved ones not being infected, and 5) confidence in fellow medical colleagues should the PCPs themselves be infected with COVID-19. The five most often utilized coping strategies by PCPs during this COVID-19 outbreak were 1) following strict personal protective measures, 2) reading about COVID-19, its transmission mechanisms and prevention, 3) engaging in relaxation activities such as exercise and prayers, 4) avoiding public spaces to minimize exposure risk, and 5) communicating with loved ones for stress relief and emotional support. Scores and subsequent rankings for each statement are presented in Table 5.

**Table 5.**
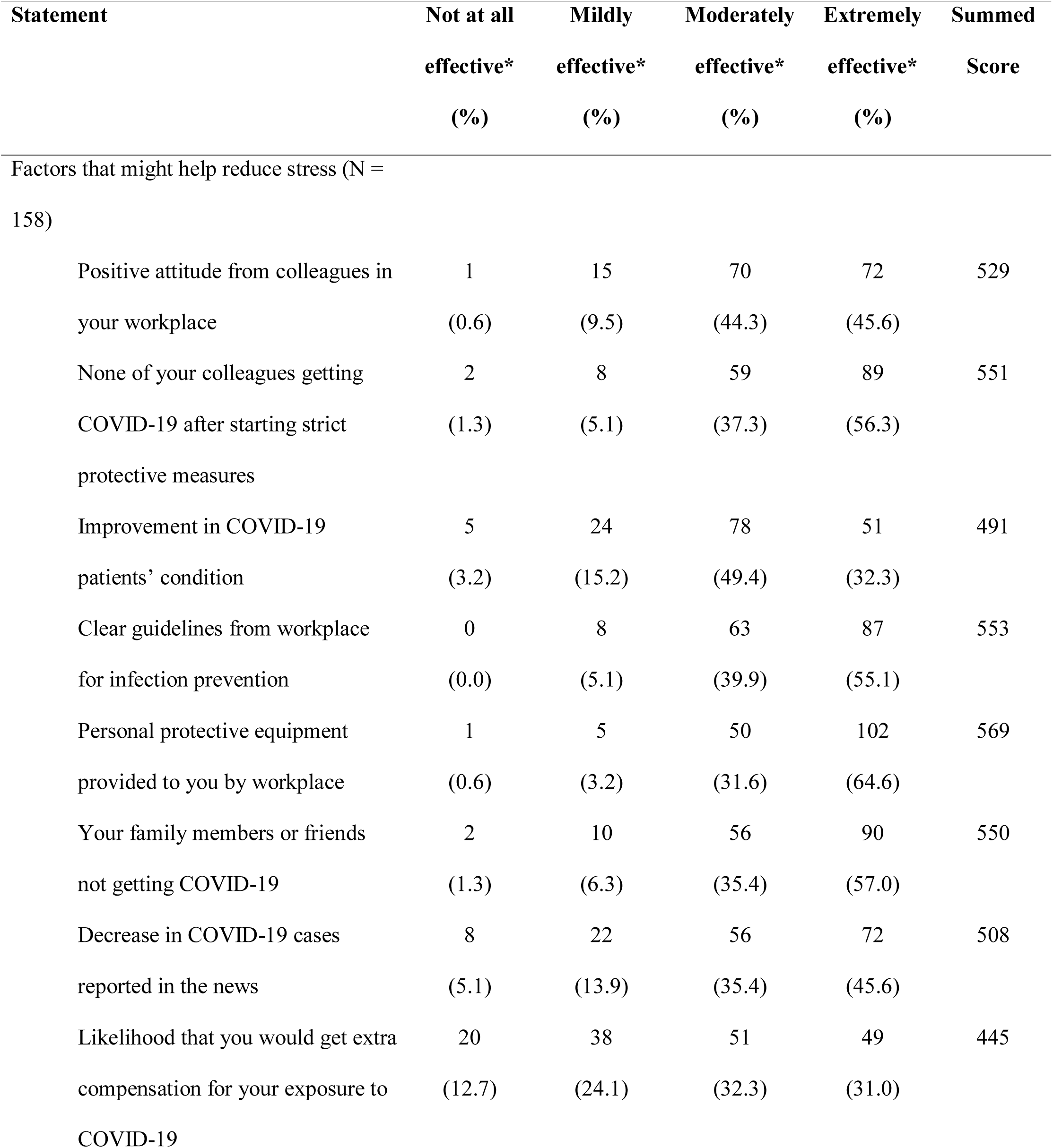
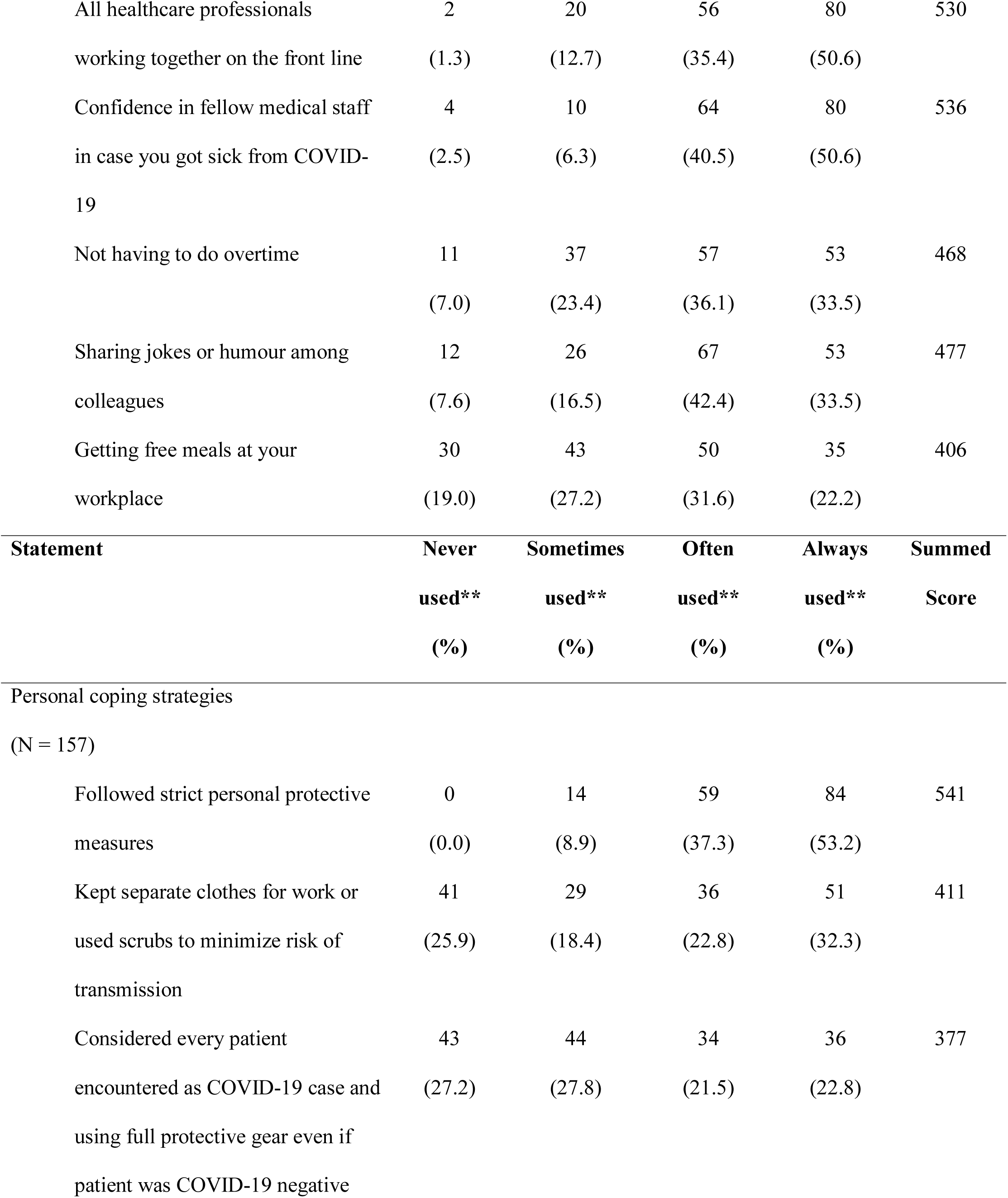
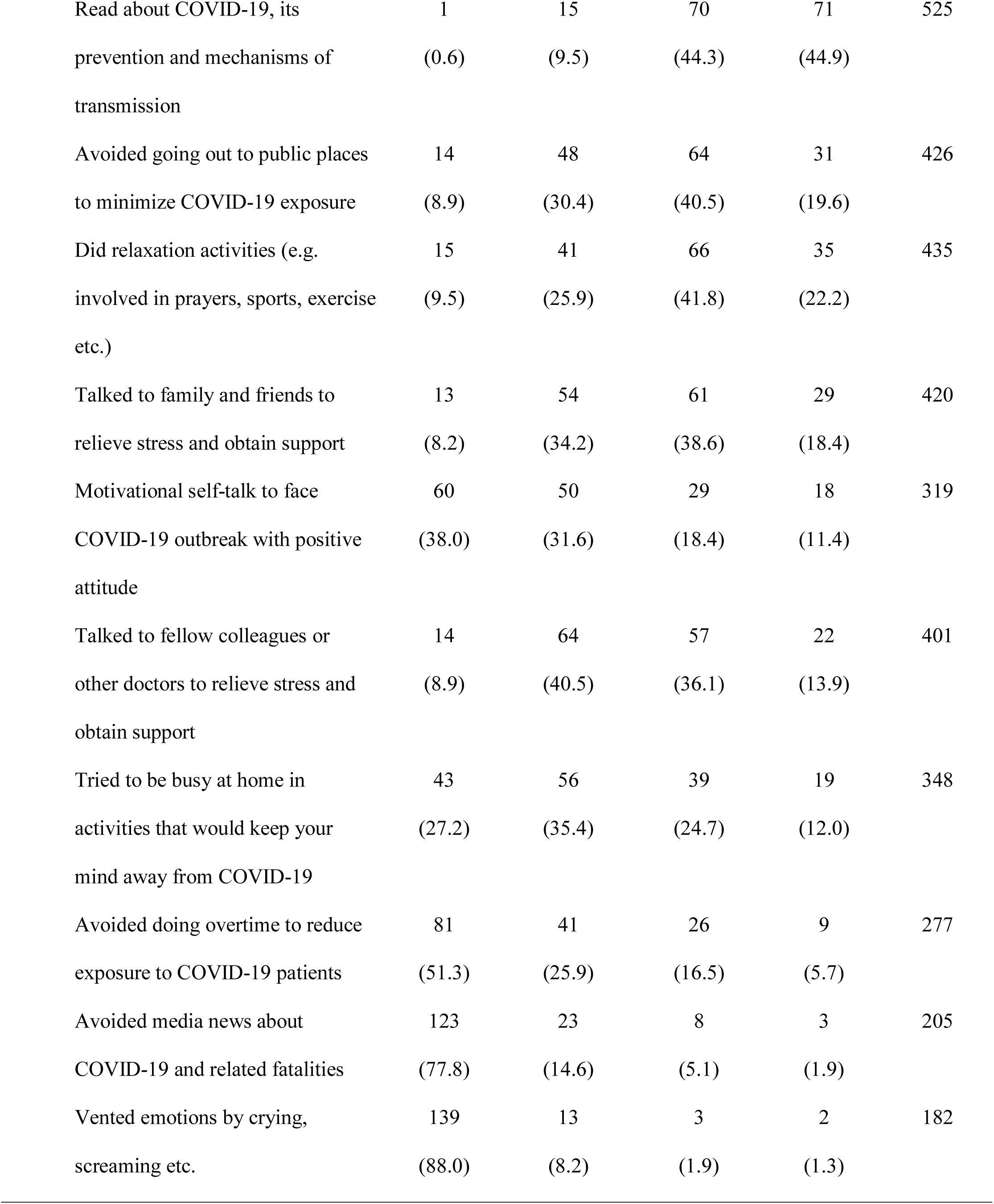
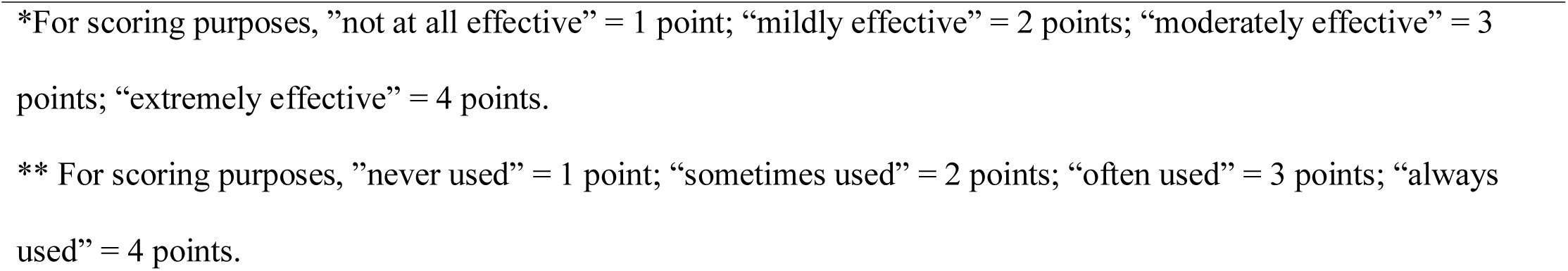
Response frequencies and summed scores for stress reduction factors and personal coping strategies.

## DISCUSSION

### PCPs’ concerns, impact on work and personal life, and outbreak preparedness

The majority of PCPs perceived themselves as being at risk of COVID-19 infection and were concerned about the risks that they could be bringing to their loved ones. Interestingly, unlike findings from similar studies conducted in Singapore during the SARS and H5N1 outbreaks, most PCPs did not feel that they or their loved ones would face social stigmatisation (2, 24)main neural network model. Moreover, the majority did not feel the need to hide the nature of their work or the risks that they could potentially face from loved ones and the public. Only a very small proportion of PCPs had considered resigning from their jobs, and the acceptance and readiness to provide medical care for COVID-19 patients was very high. PCPs differed in their opinions on colleagues resigning due to their fears, although a higher proportion (54.4%) responded with acceptance. Most seemed to feel that the COVID-19 outbreak would increase their workload and work-related stress, but would not otherwise be a cause of workplace conflict. These findings suggest a high level of ethical obligation and medical professionalism in our sample of PCPs to continue serving their communities, despite the personal and family risks incurred.

Perceived preparedness for the COVID-19 outbreak was extremely high amongst the sample. There are two possible reasons for this encouraging finding. Firstly, the Singapore Ministry of Health has gradually strengthened overall pandemic preparedness within the healthcare system since the national experiences of SARS and H1N1, which has resulted in readily available outbreak response frameworks, guidelines and protocols for both healthcare professionals and the lay public (25). The second possibility is that nearly two-thirds of the private practice PCPs in our sample were members of PHPCs. Under this scheme, their medical practices would specifically have been prepared for outbreak management with three months’ worth of PPEs provided at no cost, priority medical supplies, and standardized training on infection prevention measures (4, 26)main neural network model.

### Stress management and coping strategies among PCPs

Clear infection prevention guidelines and personal protective measures, access to information about COVID-19 and a reliable supply of PPEs were the most important factors for PCPs in coping with stress during the outbreak. This corroborates with what has been reported in SARS and MERS-CoV, where safety precautions and prompt knowledge about the disease were key motivators in incentivizing healthcare professionals to continue providing medical care through an outbreak (17, 27, 28). PCPs also rated the prevention of their medical colleagues and loved ones from being infected with COVID-19 as the next most important factor in their own stress management. This is perhaps expected given the aforementioned finding that PCPs perceived themselves as being a source of infection risk and concern to their family members due to the nature of caring for COVID-19 patients.

As it was with healthcare professionals during the SARS, H1N1 and MERS-CoV outbreaks, PCPs are likely to suffer immense emotional pressure should their colleagues and loved ones fall victim to COVID-19; considerable research has also highlighted the tendency for healthcare professionals to disregard their own physical and mental wellbeing, especially during crises (17, 27, 29). Health authorities and healthcare institutions should continue to ensure that PCPs are well provisioned not only with PPEs, but also accurate and timely information on COVID-19’s disease characteristics, transmission mechanics, as well as best practices to protect themselves, fellow medical colleagues, and – indirectly – their loved ones during this pandemic. Healthcare institutions should also seek to monitor their PCPs’ mental health and provide stress management services; studies post-SARS have cautioned that PCPs and other healthcare professionals directly involved with outbreak management are at heightened risk of psychological distress or post-traumatic stress disorder (30, 31)main neural network model. Stress management strategies could take the form of both formal and informal peer support groups, crisis counselling hotlines, or provisions for personal as well as group mindfulness exercises (29, 32)main neural network model.

### Limitations and implications for future research

As this study was designed to meet the expediencies of the ongoing COVID-19 pandemic, several key limitations must be taken into consideration. Most significantly, the generalisability of our findings is limited by the non-random sampling strategy employed and the low response rate. This was perhaps expected due to the frontline and high intensity nature of PCPs’ roles as outbreak control efforts continue to implemented in Singapore, and we recognize that our findings may only represent PCPs who were able to find the time to respond amidst their hectic schedules.

We also recognize that the quantitative descriptive nature of the present study could mean that PCPs may have had other concerns that were not included in our questionnaire. We were also unable to qualitatively probe the stress-reduction and coping strategies to understand the opinions of PCPs on how the government or healthcare institutions might be able to better support their needs.

Future studies could seek to overcome some of these limitations by implementing a more rigorous, probabilistic sampling strategy and utilizing mixed-methods designs to include qualitative interviews with PCPs. Nonetheless, the feasibility of these suggestions will inevitably depend on the progress of the COVID-19 pandemic in Singapore and around the world; social distancing measures and overwhelmed healthcare systems, for example, may prove to be immense challenges in conducting research with PCPs or other healthcare professionals.

## CONCLUSIONS

The COVID-19 pandemic seems to show no signs of any return to normalcy in healthcare within the near future, and primary care must – by necessity – remain on the frontlines of any national outbreak control strategy. We conclude by reiterating the Commission on a Global Health Risk Framework for the Future’s recommendation that primary care is inseparable with public health efforts in any pandemic, for a health system without effective and resilient primary care capabilities will lack the critical ability to quickly identify cases and mount a suitable infection prevention response (10). While PCPs continue to profess a steadfast commitment to pandemic preparedness and medical professionalism during such trying times, health systems should strive to ensure that their psychosocial and professional needs are well taken care of holistically.

## Data Availability

The datasets used and/or analysed during the current study are available from the corresponding author on reasonable request.

